# Predictors of vaccination card retention in Tamale Metropolis, Ghana

**DOI:** 10.1101/2023.09.28.23296305

**Authors:** Matthew Y. Konlan, Fuseini Mahama, Braimah B. Abubakari, Paul Konka, Benedict O. Appiah, Maxwell O. Yeboah, Peter G. Kwarteng, Porbilla O. Apea, Michael R. Adjei, Martin N. Adokiya, Oheneba Boadum, Hilarius A. K. Abiwu

## Abstract

**Background:** The home-based vaccination card (HBR) is an important health record for determining vaccination status of children during surveys, particularly in low- and middle-income countries (LMICs). However, there are limited evidence on the factors that influence its retention in Ghana. We assessed the predictors of vaccination card retention in Tamale Metropolis, Ghana

**Methods:** We conducted a cross-sectional study from 21^st^ December 2022 to 10^th^ January 2023 among children aged 0-59 months in the Tamale Metropolis. Multi-stage sampling was used to select caregivers of children aged 0-59 months for enrolment in the study. Data were collected using validated questionnaire through face-to-face interviews of caregivers. A vaccination card was retained if it was presented for physical inspection by research assistants. The factors that influence vaccination card retention were determined in a multivariate logistic regression analysis at p<0.05.

**Results:** A total of 1,566 eligible children were enrolled in this study. Vaccination card retention was 89.5%. Negative predictors of card retention included: being resident in the Nyohini (AOR=0.26; 95% CI=0.15-0.47) and Tamale Central (AOR= 0.52; 95% CI=0.30-0.92) sub-Metro areas, and children belonging to age group 24-59 months (AOR=0.40; 95% CI=0.23-0.67). On the other hand, paying for the vaccination card (AOR=5.20; 95% CI=2.98-9.04) was a positive predictor of vaccination card retention.

**Conclusion:** In this study, vaccination card retention among children aged 0-59 months was higher than national estimates. Vaccination card retention was mainly influenced by place of residence, age of child and mode of acquisition of the card such as out-of-pocket payment. There is need to design and deliver tailored messages including the importance of vaccination card retention to caregivers of children based on geographic context. Additionally, the vaccination cards should be supplied to health facilities and made readily accessible to caregivers to prevent out-of-pocket payment.

## Introduction

Routine childhood vaccination is one of the most successful public health interventions, averting four million deaths every year [1]. Aside protection against preventable diseases, vaccination brings children and their families into contact with health systems, providing an opportunity for delivery of other health services [1]. Routine immunization reduces vaccine-preventable disease incidence [2], and antimicrobial use [3], resistance [4] and associated deaths [5].

In low- and middle-income countries (LMICs), home-based vaccination cards (HBRs) are the primary means for assessing vaccination status of children during surveys [6,7]. Additionally, HBRs serve as a medium for the delivery of reminder strategies on when to bring a child for future vaccinations [8], promoting adherence to routine childhood vaccination uptake [9]. However, they are less valued, retained and used by caregivers to support healthcare decisions [10].

In Ghana, vaccination data are collected during service delivery through vaccination registers and individual HBRs. The vaccination registers are paper-based records kept in the facility and used for estimating administrative coverages. The HBRs are kept by caregivers of children for follow ups on the vaccination schedule and used as reference source for vaccination coverage surveys.

Current available evidence revealed a national vaccination card retention rate of 78.5% [11], a decline from 2014 rate of 80% [12]. Understanding factors that influence vaccination card retention can help in the design and delivery of tailored messages to improve retention. However, studies on predictors of vaccination card retention are limited in Ghana. To fill this gap, and further understand factors for non-retention of vaccination cards among caregivers, this study assessed predictors of vaccination card retention among children aged 0-59 months in Tamale Metropolis, Ghana.

## Materials and Methods

### Study design

This was a community-based analytical cross-sectional study conducted from 21^st^ December 2022 to 10^th^ January 2023 among children aged 0-59 months in the Tamale Metropolis. This study was carved out of a larger community-based cross-sectional pre- and post-COVID-19 pandemic cluster design survey of children under five years in Tamale Metropolis.

### Study area

This study was conducted in Tamale, which doubles as the administrative capital and district capital of the Northern Region and Tamale Metropolis respectively. According to the 2021 Population and Housing Census, the Metropolis has a population of 374,744: 185,051 males and 189,693 females [13]. The Metropolis is divided into four (4) health sub-Metros: Vittin, Bilpela, Nyohini and Tamale Central, with 77 health facilities including Community-based Health Planning and Services (CHPS), which render a range of services including vaccination.

### Study population

Children aged 0-59 months and their caregivers constituted the study population. Even though Ghana’ s vaccination schedule targets children from 0-18 months, we included all children less than five (5) years to compare findings of children still in the vaccination schedule and those who should have completed their schedule.

### Sample size calculation

Based on the WHO survey sample size calculator version 2 [14], we estimated a sample of 1, 512 children aged 0-59 months using the following assumptions: 95% coverage for all basic immunizations (Northern Regional target), a precision of ±5% (95% confidence interval), a design effect (DEFF) of 2, assumption of a minimum of 5 children per cluster, an intra-cluster correlation coefficient (ICC) of 1/3, assuming that 10% of eligible respondents will not be available or will decline the survey and that we would need to visit 4 households to identify an eligible child.

### Selection of respondents

Multi-stage sampling was used to select respondents. In stage one, all 63 clusters were purposively selected from the four (4) sub-Metros in Tamale. Clusters were made up of demarcated CHPS zones in the study area, and a list of all clusters and the communities under each cluster was obtained from the Tamale Metro Health Directorate. In stage two, a list of all communities in each cluster was generated and coded. Sixty-three (63) communities, one from each cluster were then selected using simple random sampling. In stage three, the initial house was selected using the central pen-spinning technique. Subsequently, houses were selected using an interval of three and two in urban and peri-urban localities respectively. In stage four, simple random sampling was used to select eligible children.

#### Eligibility criteria

All caregivers of children aged 0-59 months were included in this study. Caregivers of children above 59 months, those who had never received a vaccination card, and those who declined were excluded.

### Data collection

Data were collected using validated questionnaire by the WHO for assessing routine immunization coverage [15]. The questionnaire consisted of socio-demographic variables of eligible caregiver-child pairs, vaccination coverage using card and caregiver recall of vaccination questions. Questionnaire was deployed using the latest android version (v2021.2.4) of KoboCollect App [16] in Samsung mobile tablets. Trained research assistants collected data through face-to-face interviews with respondents. During the survey, caregivers were requested to present vaccination cards to confirm retention.

### Quality assurance

Research assistants were given a day’s training on survey objectives, methods and how to ask questions to elicit appropriate responses. Tools were pretested and modified by investigators to enhance clarity. Biodata and vaccination record pages of eligible children were taken to confirm card retention. Submitted questionnaires were checked for correctness and completeness.

### Operational definitions

A vaccination card was defined as any document given by a healthcare provider stating the vaccination status of an eligible child [17].

A caregiver is defined as an individual who provides physical and psychological care for the child.

Card was retained if a document was seen and verified by research assistants as proof of vaccination status of eligible child.

## Data analysis

Data were exported from KoboCollect into excel for cleaning before being imported in Stata Version 15.1 (Stata Corp (2017) Stata Statistical Software: Release 15. College Station, TX.) for analysis. Card retention was the outcome variable, which was determined through correct “Yes”/“No” responses to the physical inspection of a vaccination card presented for review of research assistants by caregivers as proof of vaccination status. The exposure variables were sub-Metro, sex of child, age of child, place of delivery, sex of caregiver, age of caregiver, employment status of caregiver, and educational status of respondent, relationship of child to respondent and whether card was paid for (out-of-pocket).

The predictors of vaccination card retention was assessed through a logistic regression model. The factors that showed significant association in the univariate regression analysis at p<0.05 were fitted in the multivariate regression analysis.

## Ethical Approval

This study received approval from the Navrongo Health Research Center Institutional Review Board (Approval ID: NHRCIRB495). Verbal consent was obtained from caregivers of eligible children before enrolment in the study. Study objectives, approach and rights of respondents were explained in detail and clarifying questions addressed by the research assistants.

## Results

### Sample description

Though our estimated sample size was 1512, we enrolled 1577 eligible children. However, only 1,566 were included in the data analysis after cleaning.

### Vaccination card retention

Of the 1,566 sample, 89.5% presented vaccination cards as proof of vaccination status. Of the remaining 164, misplaced (63.4%) was the predominantly reportedly reason for non-retention, followed by other reasons such as not knowing where it is being kept during the study or feeling lazy to retrieve card (28.1%), and destroyed (8.5%).

In a multivariate regression analysis, sub-Metro type, age of child and whether card was paid for or not were significant predictors of vaccination card retention. Living in Tamale Central (AOR= 0.52; 95% CI=0.30-0.92) or Nyohini (AOR= 0.26; 95% CI=0.15-0.47) sub-Metros was negatively associated with card retention. Similarly, compared to children aged 0-11 months, those aged 24-59 months had lower odds (AOR=0.40; 95% CI=0.23-0.67) of retaining their vaccination cards. However, caregivers who paid for the card had higher odds (AOR=5.20; 95% CI=2.98-9.04) of retention (Table 1).

**Table 1:** Factors associated with vaccination card retention in Tamale Metropolis, Ghana.

|  | Retention (n=1,566) |  |  |  |
| --- | --- | --- | --- | --- |
| Variable | Yes | No | COR (95% CI) | AOR (95% CI) |
| **Sub-Metro type** |  |  |  |  |
| Bilpela | 385 | 34 | Reference | Reference |
| Nyohini | 311 | 44 | 0.62(0.39-1.0) | 0.26(0.15-0.47) * |
| Tamale Central | 370 | 55 | 0.59(0.38-0.93) * | 0.52(0.30-0.92) * |
| Vittin | 336 | 31 | 0.96(0.58-1.59) | 0.93(0.48-1.83) |
| **Sex of child** |  |  |  |  |
| Female | 662 | 80 | Reference |  |
| Male | 740 | 84 | 1.06(0.77-1.47) |  |
| **Age of child(n=1532)** |  |  |  |  |
| 0-11 months | 378 | 22 | Reference | Reference |
| 23-23 months | 378 | 28 | 0.79(0.44-1.40) | 0.70(0.28-1.38) |
| 24-59 months | 646 | 80 | 0.47(0.29-0.77) * | 0.40(0.23-0.67) * |
| **Place of delivery(n=1564)** |  |  |  |  |
| Home/TBA | 95 | 6 | Reference |  |
| Privately owned facility | 45 | 2 | 1.42(0.28-7.32) |  |
| Publicly owned facility | 1260 | 156 | 0.51(0.22-1.18) |  |
| **Age of respondent** |  |  |  |  |
| <30 years | 952 | 102 | Reference |  |
| ≥30 years | 450 | 62 | 0.78(0.56-1.09) |  |
| **Sex of respondent** |  |  |  |  |
| Female | 1328 | 149 | Reference | Reference |
| Male | 74 | 15 | 0.55(0.31-0.99) * | 3.67(0.50-26.83) |
| **Educational status of respondent** |  |  |  |  |
| None | 316 | 40 | Reference |  |
| Primary | 702 | 70 | 1.27(0.84-1.91) |  |
| Secondary | 281 | 42 | 0.85(0.53-1.34) |  |
| Tertiary | 103 | 12 | 1.09(0.55-2.15) |  |
| **Employment status of respondent** |  |  |  |  |
| Employed | 420 | 53 | Reference |  |
| Unemployed | 982 | 111 | 1.12(0.79-1.58) |  |
| **Relationship of child to respondent** |  |  |  |  |
| Mother | 1336 | 145 | Reference | Reference |
| Father | 54 | 12 | 0.49(0.26-0.93) * | 0.39(0.05-2.89) |
| Other<sup>b</sup> | 12 | 7 | 0.19(0.07-0.48) * | 0.25(0.05-1.29) |
| **Pay for card (1,539)** |  |  |  |  |
| No | 829 | 132 | Reference | Reference |
| Yes | 554 | 24 | 3.68(2.34-5.75) * | 5.20(2.98-9.04) * |

<sup>b</sup>=auntie/sibling; \*p value<0.05; COR=Crude Odds Ratio; AOR=Adjusted Odds Ratio; CI=Confidence Interval; TBA=Traditional Birth Attendant

## Discussion

Our study shows a high (89.5%) vaccination card retention rate. This is similar to nationally reported rate of 80% in 2014 [12] and 78.5% in 2022 [11]. A study in Nepal [18] reported similar card retention rate of 82.2%. In contrast, lower rates of retention have been reported by other studies in Senegal (73.7%) [19], Uganda (66.0%) [20], Cameroon (29.9%) [21], Zimbabwe (68%) [22], Nigeria (63.3%, 32.6%) [23,24] and Pakistan (66.2%).

Residing in Nyohini or Tamale Central sub-Metros was negatively associated with vaccination card retention. This may be partly explained by the fact that caregivers in these sub-Metros may have low awareness level on the importance of keeping the vaccination card, as this may have not been emphasized by healthcare providers who issued cards.

Similarly, children aged 24-59 months had reduced odds of having their cards retained. Studies in Senegal [19], Uganda [20] and Pakistan [26] have reported similar findings that older children were less likely to retain vaccination cards. The possible reason for this is that caregivers don’t understand the importance of keeping the card after completing the vaccination schedule.

Caregivers who paid for the vaccination card had increased odds of retaining it. The possible reason is that caregivers who pay for the vaccination cards recognize its value and are more likely to take good care of them. On the other hand, they may want to avoid paying for replacement in event of a loss. However, sale of vaccination cards is against the vaccination policy in Ghana, even in times of shortages, when health facilities print copies to support service delivery. It is expected that the cost of making copies of the vaccination card be borne by the health facilities without transfer to service users.

### Strengths and limitations of the study

This study used a relatively large sample; therefore, the findings can be interpreted in wider context. Additionally, the study used an objective measure through physical inspection to assess card retention. However, findings of this study should be interpreted in the light of the fact that as cross-sectional study, it difficult to establish causality between exposures and vaccination card retention. Additionally, the failure of some of the caregivers to present cards for inspection does not mean that the children do not have them.

## Conclusions

Our study found high vaccination card retention rate. Sub-Metro, age of child and payment for card were significantly associated with vaccination card retention.

Health care providers in the Nyohini and Tamale Central sub-Metros should endeavor to intensify education of caregivers on the importance vaccination cards and the need for safe keeping. Additionally, caregivers of older children should be educated about the importance of keeping cards after the child has completed vaccination schedule. Given that payment for card improves retention, vaccination cards should be supplied to health facilities and made readily accessible to caregivers to prevent out-of-pocket payment.

## Acknowledgement

We thank the caregivers of children aged 0-59 months in the study area and the field officers for their dedicated service during fieldwork. We acknowledge funding support from the Northern Regional Health Directorate and the Sabin Vaccine Institute Boost Community.

## Data Availability statement

All relevant data are included in the manuscript and will be provided on reasonable request.

## Authors’ contributions

Conceptualization: Matthew Yosah Konlan, Dr. Hilarius Asiwome Kosi Abiwu, Fuseini Mahama, Dr. Braimah Baba Abubakari.

Data curation: Matthew Yosah Konlan, Dr. Hilarius Asiwome Kosi Abiwu.

Formal analysis: Matthew Yosah Konlan.

Methodology: Matthew Yosah Konlan, Dr. Hilarius Asiwome Kosi Abiwu, Dr. Braimah Baba Abubakari, Fuseini Mahama, Paul Konka, Benedict Ofori Appiah, Maxwell Oduro Yeboah, Dr. Peter Gyamfi Kwarteng, Porbilla Ofosu Dr. Oheneba Boadum, Dr. Michael Rockson Adjei, Professor Martin Nyaaba Adokiya.

Supervision: Matthew Yosah Konlan, Fuseini Mahama, Dr. Hilarius Asiwome Kosi Abiwu, Dr. Braimah Baba Abubakari.

Writing – original draft: Matthew Yosah Konlan.

Writing – review & editing: Dr. Hilarius Asiwome Kosi Abiwu, Dr. Braimah Baba Abubakari, Fuseini Mahama, Paul Konka, Benedict Ofori Appiah, Maxwell Oduro Yeboah, Dr. Peter Gyamfi Kwarteng, Porbilla Ofosu Apea, Dr. Oheneba Boadum, Dr. Michael Rockson Adjei, Professor Martin Nyaaba Adokiya

## List of abbreviations

AOR: adjusted odds ratio
CHPS: community-based health planning and services
CI: confidence interval
COR: crude odds ratio
DEFF: design effect
HBR: home-based vaccination cards
ICC: intra-cluster coefficient
LMICs: low- and middle-income countries
TBA: traditional birth attendant

